# Epidemiology of vascular access for outpatient hemodialysis procedures and renal transplantation in Brazil between 2008 and 2022: A cross-sectional nationwide analysis

**DOI:** 10.1101/2025.01.15.25320581

**Authors:** Bruno Jeronimo Ponte, Carolina Carvalho Jansen Sorbello, Lucas Redivo Basani, Marcelo Fiorelli Alexandrino da Silva, Marcelo Passos Teivelis, Sergio Kuzniec, Nelson Wolosker

## Abstract

3.

**BACKGROUND AND HYPOTHESIS:** Chronic kidney disease (CKD) represents a significant global public health challenge, affecting up to 13.4% of the population and ranking as the third fastest-growing cause of mortality. Progression of CKD to end stage kidney disease (ESKD) is the initial milestone for renal replacement therapy, which includes hemodialysis and renal transplantation, the latter marking the final therapy for ESKD patients. Arteriovenous fistulas (AVFs) are preferred for hemodialysis due to lower infection risks and superior patency compared to long-staying tunneled catheters (LSTC). Despite the importance of vascular access and transplantation, nationwide data correlating their utilization over time remains scarce. This study aimed to analyze outpatient hemodialysis vascular access procedures in Brazilian public health system from 2008 to 2022, comparing the temporal trends with renal transplantation.

**METHODS:** This study employed a cross-sectional, population-based analysis of publicly available data pertaining outpatient vascular access procedures for hemodialysis and renal transplantation in the brazilian public health system between 2008 and 2022. Linear regression and correlation between both were then carried out.

**RESULTS:** Between 2008 and 2022, 937,739 procedures were realized, including vascular access confection, ending procedures and renal transplants. Arteriovenous fistulas accounted for the majority of procedures (55%), followed by LSTC (34%), renal transplants in cadaveric donors (5.6%), catheter removals (2.2%), renal transplants with living donors (1.6%) and fistula ligation (1.4%). The increase in AVF creation was not proportional to the growth of the hemodialysis population *(p<0.001)*, in contrast to catheter implants, which increased substantially over the study period (*p<0.001*). Vascular access-ending procedures demonstrated a significant correlation with renal transplants *(p<0.001)*.

**CONCLUSION:** In Brazil, the number of LSTC for hemodialysis has increased over the years, while the number of AVFs significantly decreased. Additionally access-ending procedures, such as fistula ligation and catheter removals, have risen annually since 2010 and demonstrated a correlation with renal transplantation.

Key Points:

## 4. INTRODUCTION

Chronic kidney disease (CKD) is a significant global public health issue (1), affecting approximately 10% of the world’s population, with some studies suggesting a prevalence as high as 13.4%. (2,3) It is currently the third fastest-growing cause of mortality and is projected to become the fifth leading cause of years of life lost by 2040. (4) As CKD progresses to end-stage kidney disease (ESKD), treatments such as hemodialysis and renal transplantation become necessary (1), with approximately 4 million people estimated to need renal replacement therapy (RRT) worldwide. (1,4)

CKD not only impacts individuals by reducing their quality of life and productive capacity and increasing the risk of death (5) but also creates financial pressure on health systems due to the costs of hospitalization, surgical interventions, and loss of work activities.(6,7)

To improve CKD patients’ survival and quality of life, it is crucial to focus on vascular accesses, such as arteriovenous fistulas (AVFs) for hemodialysis and renal transplantation. AVFs are preferred for hemodialysis as they present a lower risk of infection and greater patency than long-staying tunneled catheters (LSTC). (8–10) Additionally, renal transplantation significantly benefits CKD patients and those on RRT. (9)

Understanding the temporal trends of vascular access procedures and renal transplantation is crucial for establishing effective public health policies. Several hemodialysis censuses are carried out worldwide, establishing the number of patients on hemodialysis, which vascular access is used, and how many patients are referred for renal transplantation. (11–13) However, to date, there have been no nationwide data demonstrating the number of AVFs or LSTC implants performed over the years.

In Brazil, with a population of around 215 million, the Unified Health System (SUS) was created in 1988 to provide universal access to healthcare financed by taxes. Despite this, around 25% of individuals use the supplementary health system as their main healthcare service, while 75% rely exclusively on the public system for medical services. (14,15)

The goal of this study was to analyze all outpatient procedures related to hemodialysis vascular access in the Brazilian public health system from 2008 to 2022. It also aimed to investigate the relationship between creating AVFs, LSTC implants, AVF ligations, LSTC removals, and renal transplants.

## 5. METHODS

This was a cross-sectional and retrospective design study in which we analyzed data obtained from DATASUS, a Ministry of Health platform that gathers information on hospitalizations and outpatient procedures financed by SUS. The institution’s Research Ethics Committee approved the study. This platform is public and feeds unidentified data, which is why there was no need to apply an informed consent form.

### 5.1. Period analyzed, data source, and extraction

This retrospective study analyzed the available data on LSTC for hemodialysis implantation and removal, AVF confection and ligation, and renal transplantation procedures performed in Brazil’s public health network between 2008 and 2022. The data was obtained from DATASUS, a digital platform of the Unified Health System that offers open data on procedures carried out in public hospitals accredited by the system. This accreditation is mandatory for the government to reimburse institutions.

For this study, only codes referring to outpatient procedures were collected, and inpatient settings were not included.

Data were extracted from this platform using an automated extraction method developed by the institution’s IT service in Python (v. 2.7.13; Beaverton, OR, USA), using the Windows 10 operating system. Field selection on the DATASUS platform and subsequent table adjustment were carried out using Selenium WebDriver (v. 3.1.8; Selenium HQ) and Pandas (v. 2.7.13; Lambda Foundry, Inc. and PyData Development Team, NY, USA).

### 5.2. Selection of procedures on the platform

Data was selected and collected for the following coded procedures according to the Brazilian public health system’s coding.

#### 5.2.1. Selection of Catheter implantation and removal for Hemodialysis

Removal of semi or fully implantable long-term catheter (04.06.02.062-0); Long-stay Catheter Implantation for Hemodialysis (04.18.01.004-8); Long-stay Catheter Implantation for Hemodialysis (07.02.10.001-3);

#### 5.2.2. Selection of Arteriovenous fistula confection and ligation for Hemodialysis

Arteriovenous fistula confection for hemodialysis (04.06.02.008-6); Autologous Arteriovenous fistula confection (04.180.100-21); Prosthetic Arteriovenous fistula confection (04.18.01.001-3); Arteriovenous fistula ligation (04.18.02.002-7).

#### 5.2.3. Selection of Renal transplantation procedures

Renal transplantation – Living donor (05.05.02.009-2); Renal transplantation – Cadaveric donor (05.05.02.010-6).

### 5.3. Population Analysis

For the procedures, demographic data (distribution by region of the country) and data on reimbursement amounts passed on to institutions for the surgeries performed were collected. The data was compiled in .cvs format and organized into tables using Microsoft Office Excel 2016 (Redmond, WA, USA).

Population data referring to the Brazilian population, which exclusively uses SUS and hemodialysis patients, were used as denominators for the correlation analysis between the procedures under study. The population samples were collected from the Brazilian Institute of Geography and Statistics (IBGE) database and the Brazilian Dialysis Census and are described in Table 1. Of the group of dialysis patients, around 95% were on hemodialysis, while 5% were on peritoneal dialysis. The total sample of patients under dialysis was chosen as the basis for the epidemiological calculations. Between 2008 and 2022, the number of patients on dialysis increased by 76.7%. In contrast, the Brazilian population grew by 13.3%, while the percentage of the population that relies exclusively on the SUS remained relatively stable, increasing from 74.9% to 75%.

**Table 1.** Estimated total population and dialysis patients in Brazil between 2008 to 2022.

| Year | Dialysis patients<sup>1</sup> | (%) | Brazilian population<sup>2</sup> | Brazilian population under public health services exclusively |
| --- | --- | --- | --- | --- |
| 2008 | 87,044 | 0.05% | 189,612,814 | 142,209,611 |
| 2009 | 77,589 | 0.04% | 191,507,554 | 143,630,666 |
| 2010 | 92,091 | 0.05% | 194,890,682 | 146,168,012 |
| 2011 | 91,314 | 0.05% | 196,603,732 | 147,452,799 |
| 2012 | 97,586 | 0.05% | 198,314,934 | 148,736,201 |
| 2013 | 100,397 | 0.05% | 200,004,188 | 150,003,141 |
| 2014 | 112,004 | 0.06% | 201,717,541 | 151,288,156 |
| 2015 | 111,303 | 0.05% | 203,475,683 | 152,606,762 |
| 2016 | 122,825 | 0.06% | 205,156,587 | 153,867,440 |
| 2017 | 126,583 | 0.06% | 206,804,741 | 155,103,556 |
| 2018 | 133,464 | 0.06% | 208,494,900 | 156,371,175 |
| 2019 | 139,691 | 0.07% | 210,147,125 | 157,610,344 |
| 2020 | 144,779 | 0.07% | 211,755,692 | 158,816,769 |
| 2021 | 148,363 | 0.07% | 213,317,639 | 159,988,229 |
| 2022 | 153,831 | 0.07% | 214,828,540 | 161,121,405 |

1: *Brazilian Dialysis Census 2022 (around 95% under hemodialysis and 5% under peritoneal dialysis)*; 2: *Brazilian Institute for Geography and Statistics (IBGE)*

### 5.4. Data Analysis Linearity

Initially, an analysis of the general data of the procedures under study was carried out. The procedures performed on AVFs, catheter implants, and renal transplants were then respectively analyzed.

The order described in the results and discussion is programmatic to maintain linearity, since many correlations were made between the procedures under study and the populations analyzed.

### 5.5. Statistical Analysis and Ethics Committee approval

For this study, we analyzed the number of hemodialysis catheter-related procedures (implantation and removal), AVFs confection and ligation, and renal transplantation procedures (from living or cadaveric donors) by the years 2008 to 2022.

We analyzed the relation between the number of procedures performed each year and by region and correlated them with the Brazilian population on hemodialysis from 2008 to 2023. The statistical analysis was conducted using SPSS 20.0 for Windows (IBM Corp, Armonk, NY). Linear regression was employed to analyze the evolution of catheters, fistulas, and renal transplantation over the years. The Spearman correlation was used to compare the relation between each of the procedures analyzed. A value of p <0.001 was considered statistically significant for all tests.

The data was obtained anonymously through this platform, so a consent form was unnecessary. The study protocol was approved by the institution’s research ethics committee.

## 6. RESULTS

### 6.1. Total procedures

Over the 15 years studied, 937,739 hemodialysis access-related procedures and renal transplantations were performed (Table 2). The most frequently performed procedure was the confection of AVFs (55.02%). There has been an increase in the number of procedures over the years, except for living donor renal transplants, which demonstrated a reduction.

**Table 2.** Total procedures performed between 2008 and 2022.

| Year | AVF | AVF<br>Ligation | Catheter | Catheter<br>Removal | RT –<br>Cadaveric<br>donor | RT –<br>Live<br>donor | Total |
| --- | --- | --- | --- | --- | --- | --- | --- |
| **2008** | 26,402 | 344 | 6363 | - | 1712 | 1429 | 36,250 |
| **2009** | 28,754 | 355 | 7167 | - | 2336 | 1469 | 40,081 |
| **2010** | 29,617 | 448 | 8269 | 117 | 2516 | 1335 | 42,302 |
| **2011** | 31,019 | 497 | 9705 | 315 | 2973 | 1457 | 45,966 |
| **2012** | 33,138 | 594 | 12,470 | 637 | 3421 | 1219 | 51,479 |
| **2013** | 33,603 | 599 | 14,731 | 1121 | 3542 | 1129 | 54,725 |
| **2014** | 33,848 | 847 | 17,740 | 1211 | 3788 | 1145 | 58,579 |
| **2015** | 34,518 | 898 | 19,449 | 1477 | 3895 | 963 | 61,200 |
| **2016** | 34,567 | 922 | 21,075 | 1520 | 3900 | 966 | 62,950 |
| **2017** | 35,976 | 1057 | 22,499 | 2137 | 4240 | 896 | 66,805 |
| **2018** | 37,934 | 1176 | 26,260 | 2065 | 4399 | 797 | 72,631 |
| **2019** | 40,366 | 1389 | 31,347 | 2592 | 4617 | 857 | 81,168 |
| **2020** | 38,489 | 1632 | 38,647 | 2158 | 3864 | 331 | 85,121 |
| **2021** | 38,389 | 1480 | 40,161 | 2408 | 3641 | 443 | 86,522 |
| **2022** | 39,500 | 1488 | 43,547 | 2944 | 3900 | 581 | 91,960 |
| **Total** | **516,120** | **13,726** | **319,430** | **20,702** | **52,744** | **15,017** | **937,739** |

*AVF: Arteriovenous fistulae; RT: Renal transplant*

### 6.2. Arteriovenous fistulae confection

A total of 516,120 fistulas were performed, with an annual average of 34,408 per year (SD ± 4157). The number of AVFs performed per 1000 dialysis patients is shown in Figure 1. Although the total number of AVF procedures has increased over the years, this growth has not been proportional to the growth of the hemodialysis population, with a linear decrease over the years *(p<0.001)*.

**Figure 1.**
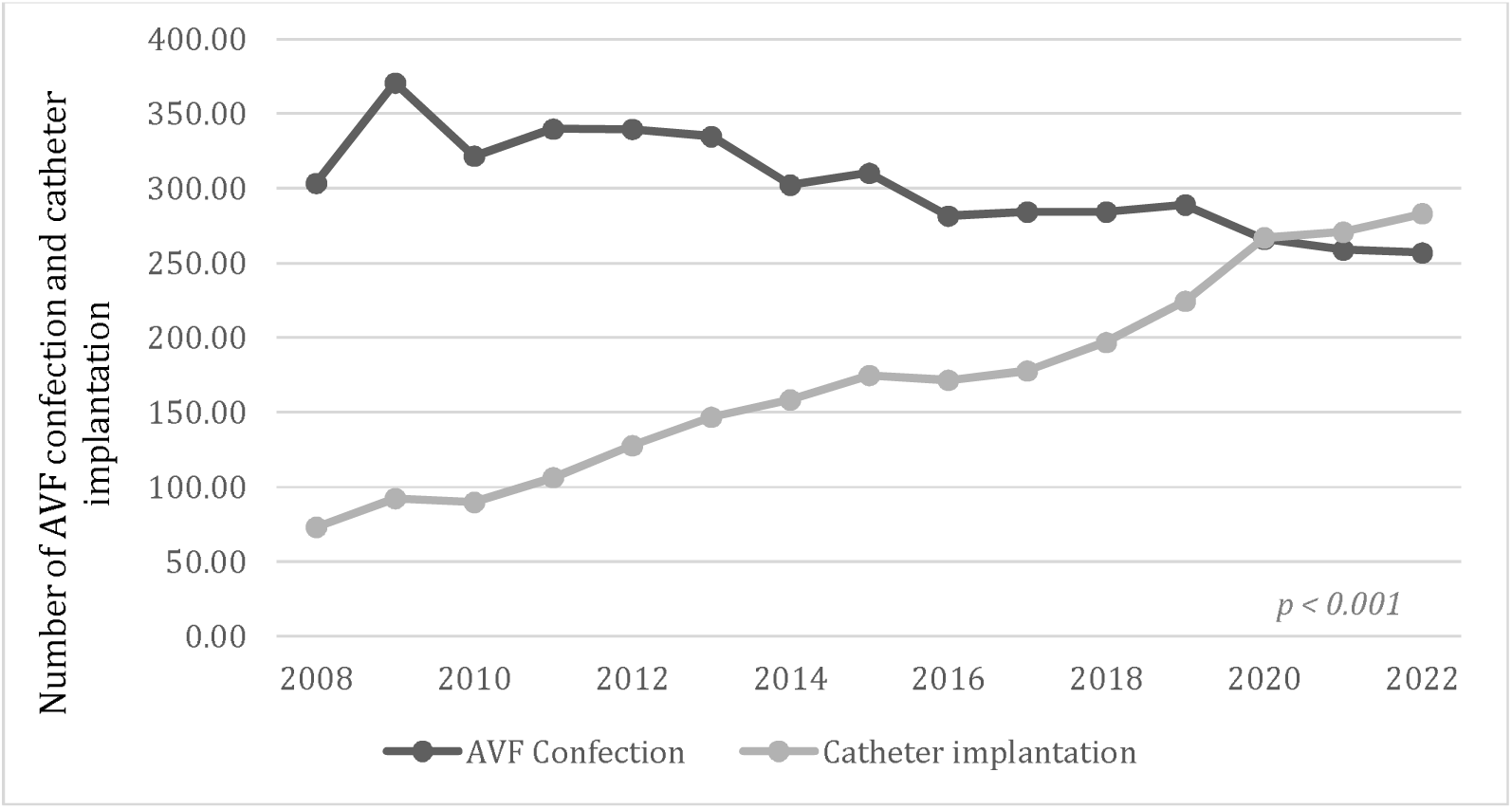
Number of AVF confection and catheter implantation per 1000 dialysis patients over the years.

### 6.3. Catheter implantation

The total number of catheters implanted was 319,430, with an annual average of 21,295 catheters per year (SD ± 12,372). The number of long-term catheter implantation per 1000 dialysis patients is demonstrated in figure 1. The increase in the number of catheters implanted correlated positively with the increase in the hemodialysis population over the years (*p <0.001*).

### 6.4. Renal transplantation

A total of 52,744 deceased donor transplants were carried out, with an annual average of 3516 (SD±806). Living donor transplants accounted for 15,017 procedures, with a yearly average of 1001 (SD±360). Renal transplantation performed per dialysis patients over the years are illustrated in Figure 2. Concerning deceased donors, there has been stability in the number of transplants compared to the dialysis population over the years despite no significant correlation being found (*p 0.935*). However, between 2019 and 2020, there was a significant drop, with a progressive increase in procedures after 2021. When analyzing the progression of living donor renal transplants, there was a significant drop in the number of procedures over the years *(p < 0.001)*.

**Figure 2.**
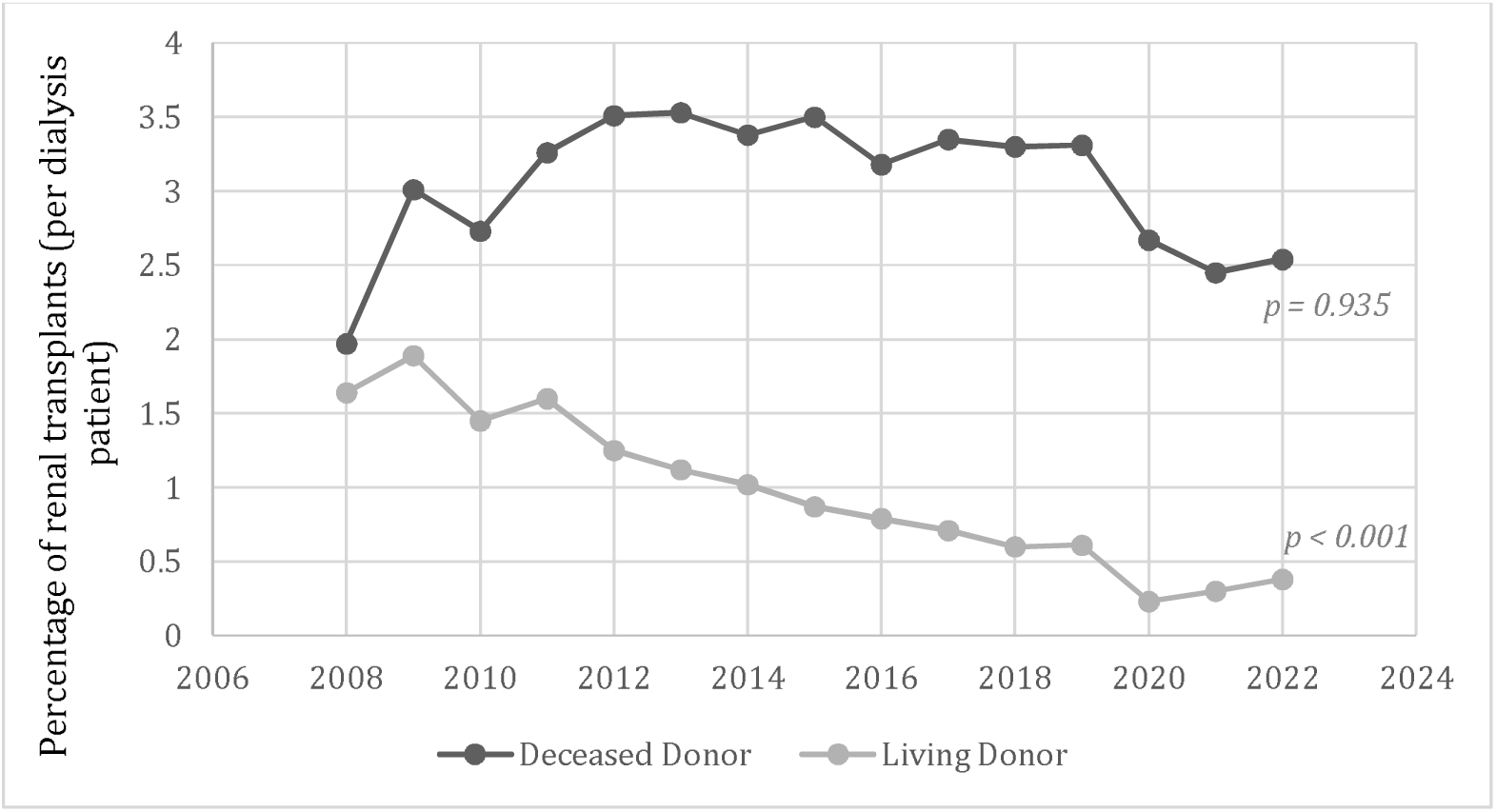
Renal transplants in proportion to the dialysis population over the years.

### 6.5. Access-ending procedures

When analyzing access-ending procedures, such as AVF ligation and catheter removal, we obtained the relation of procedures based on the number of AVFs created and catheters implanted. total of 1726 AVFs were ligated, with an annual average of 915 (SD ±440) procedures. An average of 25.56 ligatures per 1000 fistulas created was found over the years. (figure 3). There was a linear progression in the number of fistula ligations compared to the number of AVFs created (*p<0.001)*.

**Figure 3.**
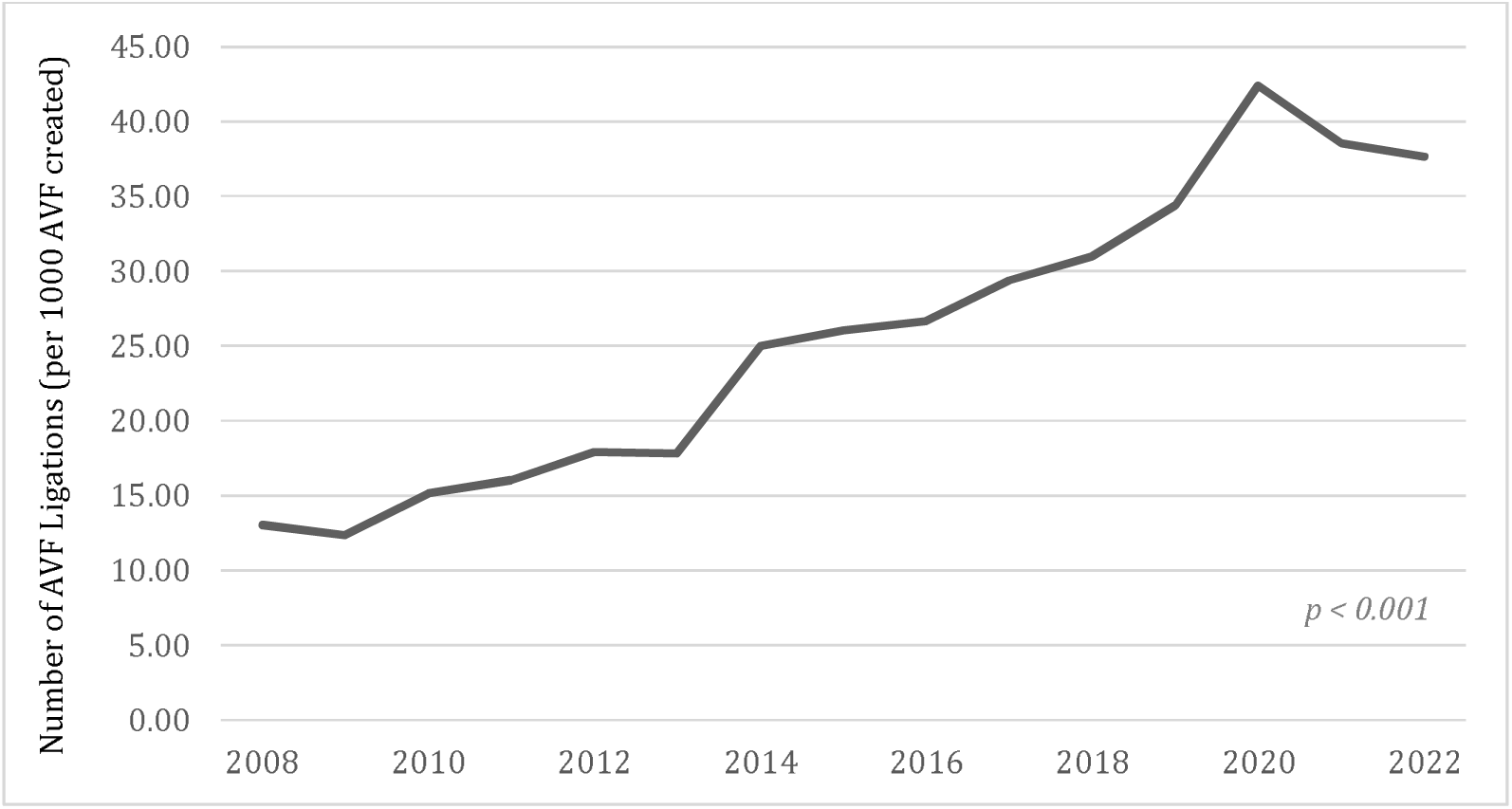
Number of AVF ligations per 1000 AVF confection over the years. Linear regression (p<0,001) AVF: Arteriovenous fistula.

A total of 20,702 catheters were removed, with an annual average of 1592 (SD±886) procedures. The average of catheter removal per 1000 implants was 63.83 over the years. The relation between catheter removal and implants are illustrated in Figure 4.

**Figure 4.**
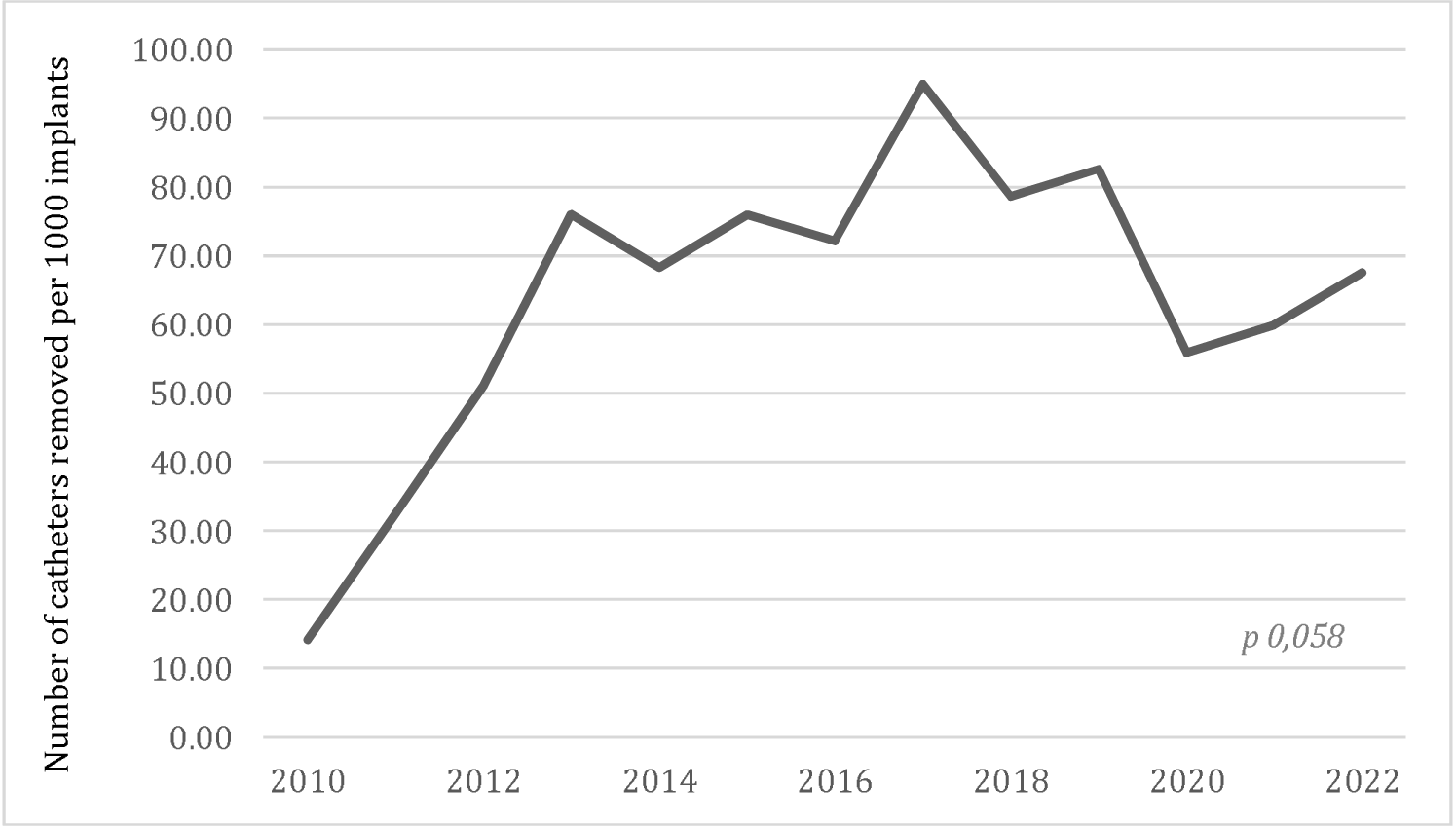
Number of catheters removement per 1000 implants over the years. Linear regression (p 0,058)

When compared with renal transplants, AVF ligation procedures showed a significant correlation with deceased and living donor transplants *(p<0.001).* Catheter removals also significantly correlated with deceased and living donor transplants *(p 0.003 and p<0.001, respectively)*.

The number of catheter implants, removals, and their costs for each Brazilian region are described in Table 3. The total reimbursed throughout the years was U$ 30,997,946.76. The overall average amount paid per procedure was U$ 91.14.

**Table 3.** Catheter Implants, removals and reimbursements by Brazilian Region.

| Region | Catheter Implant | (%) | Catheter Removal | (%) | Total of Procedures | Cost (U\$) | Mean (U\$) |
| --- | --- | --- | --- | --- | --- | --- | --- |
| North | 12,066 | 3.78% | 897 | 4.33% | 12,963 | 1,134,569.67 | 87.52 |
| Northeast | 63,928 | 20.01% | 3412 | 16.48% | 67,340 | 6,262,804.49 | 93.00 |
| Southeast | 200,212 | 62.68% | 10,524 | 50.84% | 210,736 | 19,410,455.05 | 92.11 |
| South | 33,321 | 10.43% | 5629 | 27.19% | 38,950 | 3,242,797.60 | 83.26 |
| Center-west | 9903 | 3.10% | 240 | 1.16% | 10,143 | 947,319.95 | 93.40 |
| Total | 319,430 | 100% | 20,702 | 100% | 340,132 | 30,997,946.76 | 91.14 |

## 7. DISCUSSION

### GENERAL ANALYSIS

Between 2008 and 2022, a total of 937,739 surgical procedures related to hemodialysis access and renal transplants were performed. To our knowledge, this represents the largest sample ever studied for analyzing the creation and ligation of AVFs, as well as the implantation and removal of long-term catheters for hemodialysis and renal transplants in a single country.

This study reflects the reality of the Brazilian population that utilizes the public health system (SUS) and is receiving treatment for kidney failure requiring hemodialysis. According to the Brazilian Dialysis Census, around 153,831 individuals were undergoing dialysis in 2022, of which around 95% were on hemodialysis and 5% were peritoneal dialysis patients. (12) About 75% of the Brazilian population rely exclusively on public health services (SUS), corresponding to around 165 million people, based on 2022 data. (14) Unfortunately, it was not possible to obtain information on the various modalities of hemodialysis access provided by the supplementary health service since centralized and public data for this population was unavailable. (14,15)

According to the Brazilian Dialysis Census of 2008, the estimated number of individuals on dialysis in Brazil was 87.044. Since then, there has been a gradual increase in the dialysis population. By 2022, the number rose by 76%, reaching approximately 153,831 individuals. (12) In addition, in 2022 there was an increase in the prevalence of patients using long-term catheters from 6% in 2013 to 15.3% in 2021, and a drop in the use of AVFs from 73.9% to 68%, in the same period. The annual mortality of the dialysis population in 2022 was estimated at 17.1%. (12)

### TRENDS REGARDING ARTERIOVENOUS FISTULA

Between 2008 and 2022, a total of 516.120 new AVF were created. We collected this data through three procedure codes linked to AVFs: those involving PTFE grafts, autologous grafts, and fistulae that were not specified for hemodialysis. When we stratified the procedures, we observed that 35,546 belonged to the PTFE fistula group, 17,016 were autologous, and 463,558 did not specify the type of fistula. This information indicates that most procedures did not specify the type of fistula. This lack of specification may create a bias in filling out the procedure forms, which hinders a complete analysis, preventing us from determining which kind of fistula is the most commonly performed in the country.

Furthermore, we know that the patency of arteriovenous fistulas can vary in the literature. (16). It is known that, over the years, it is common for patients to need several fistulas due to problems with maturation, patency and the need for interventions. (17) Factors such as the patient’s age, state of health, and type of fistula significantly influence this need. (16,18) The data analyzed in our study did not allow an individual analysis of the patients who underwent surgery, and it was not possible to establish how many fistulas were performed on a single patient

The main guidelines for hemodialysis access recommend creating autologous arteriovenous fistulae for patients whenever possible. (19)This approach is preferred because it offers a longer patency duration and a lower risk of long-term complications. (8,20) In addition, dialysis patients report improved quality of life when using AVF compared to using LTSC. (21)

There was an annual increase in the number of AVFs. However, compared with the significant increase in the hemodialysis population, there has been a proportional decrease in the number of AVFs per hemodialysis patient. When comparing with North American data, between 2013 and 2021 in the United States, there was a drop in patients starting hemodialysis under AVF from 17% to 12.2%, corroborating the reduction in our sample. (22) The reduction in AVFs can also be attributed to challenges in establishing quality vascular accesses, especially in vulnerable populations, particularly the elderly.(23,24) This contributes to greater dependence on other types of access, which can impact the quality of treatment and complication rates.(25)

Fistula ligation has also increased progressively over the years. From 2008 to 2022, for every 1000 fistulas created, 26.56 were ligated on an outpatient basis. A limitation of this analysis is that we do not have access to the reason the fistula was ligated. Possible causes for fistula ligation include infection, aesthetics, heart failure, steal syndrome, loss of fistula function, recovery of kidney function, and after renal transplantation. (22) But, considering that the analysis was based on outpatient procedures, we consider it unlikely that the ligatures were performed in the context of an infection or other emergency scenario, such as a rupture.

There is still no consensus in the literature regarding the ligation of fistulas after renal transplantation. The procedure’s benefits in this population include a reduction in ventricular overload due to changes in afterload, the risk of bleeding from the AVF, and aesthetics for the patient. (26) The controversy exists because patients undergoing renal transplantation may still need hemodialysis, and maintaining the fistula allows for a less invasive return to hemodialysis, thereby avoiding the need for new procedures to access hemodialysis. (26,27) In our analysis, AVF ligations were significantly correlated with renal transplant procedures (p<0.001), demonstrating a plausible preference for ligation of the AVF after renal transplantation in the various Brazilian services.

### TRENDS REGARDING LONG STAY TUNNELED CATHETERS FOR HEMODIALYSIS

In accordance to what we observed with fistulas, the number of catheter implants has gradually increased over the years, which is in line with the growing hemodialysis population. According to US data, between 2013 and 2021, there was an increase of approximately 13.7% in patients starting hemodialysis with LSTC. (22) This information supports the data found in our sample, demonstrating a greater trend toward catheter implantation in hemodialysis patients over recent years.

To date, there are no studies in the literature that indicate the number of implants for long-term catheters for hemodialysis. During our data collection, we noticed a temporal trend towards more catheters being implanted in dialysis patients, which coincided with a proportional decline in the number of AVFs being created, demonstrating a trade-off between the types of vascular access for hemodialysis.

Despite this increase, LSTC are routes for hemodialysis that tend complicate more than AVFs, with the possibility of infection, tip migration, lumen thrombosis with dysfunction and thrombosis of central venous sites. (28,29) When it comes to removing catheters many reasons can justify the removal of an LSTC, including dysfunction, infection, recovery of renal function, maturation of the AVF, and termination of hemodialysis after renal transplantation. (26,30)

Due to the scope of the study and the data collected, the reasons for catheter implantation and removal were not established. As a result, we did not recognize the reasons for implanting catheters on an outpatient basis, nor their removal.

Analyzing the data on catheter removal shows that for every 1000 LSTCs implanted, 63.83 catheters are removed. In addition, there was a correlation between the number of catheter implants, AVFs, and renal transplants in living donors in our sample. This leads us to believe that more catheters are removed as the number of more definitive and long-lasting accesses, such as AVFs, increase. Also, as patients are transplanted, more hemodialysis catheters are removed.

### TRENDS REGARDING RENAL TRANSPLANTATION

The data on kidney transplant projections aligns with the findings reported by the Brazilian Organ Transplant Association (ABTO), which annually documents the prevalence of transplants in the country. (31) There has been an absolute increase in the number of renal transplants, predominantly at the expense of cadaveric donors, with a drop in the number of transplants using living donors.

Compared to the international data, we observe that the number of living donor transplants in Brazil has decreased yearly. In contrast, in the United States and Europe, the number of living-donor transplants has remained stable and is on the rise. (32,33) Furthermore, Brazil’s number of cadaveric donors’ renal transplantation per hemodialysis patient has shown a tendency towards stability.

We noticed a drop during 2020 and 2021, followed by an upward trend over the last two years. This drop is compatible with the global scenario during the COVID-19 epidemic, when the number of elective surgical procedures, including transplants, fell significantly. (19–22) The subsequent years indicate a projected increase in the number of cadaveric donors, also corroborating global literature. (32–35)

The correlation in our country between the number of renal transplants and vascular access closure procedures is remarkable. As the number of renal transplants has risen, there has also been a linear increase in the number of AVF ligations and catheter removals.

### LIMITATIONS

As with any database study, the accuracy of the results is susceptible to errors inherent in coding or data entry. Since the data is anonymous and the analysis is retrospective, it was not possible to follow up patients longitudinally. Consequently, this study did not access data related to patients’ comorbidities and clinical outcomes. For this reason, information on procedures per patient is not available, considering that the same patient can have more than one AVF, more than one catheter implanted or even be retransplanted.

Regarding vascular access procedures, this study only analyzed procedure codes for patients treated on an outpatient basis and did not include inpatient cases. Consequently, the total values were underestimated since these cases were not included in the analysis.

Also, this study only analyzed the population that exclusively uses the public health service, which represents the majority of the Brazilian population, but it could lead to an underestimated analysis of surgeries related to vascular access for hemodialysis and renal transplantation.

## 8. CONCLUSION

In Brazil, the number of long-term catheters implanted for hemodialysis has increased over the years, while the number of arteriovenous fistulas significantly decreased. Additionally access-ending procedures, such as fistula ligation and catheter removals, have risen annually since 2010.

Regarding transplants procedures, there has been a gradual decline in the number of transplants performed, with the COVID-19 pandemic a significant drop in these activities. However, after 2021, we have begun to see an upward trend in transplant number.

## Data Availability

All data produced in the present study are available upon reasonable request to the authors

